# Impact of multiple sclerosis disease-modifying therapies on SARS-CoV-2 vaccine-induced antibody and T cell immunity

**DOI:** 10.1101/2021.09.10.21262933

**Authors:** Joseph J. Sabatino, Kristen Mittl, William Rowles, Kira Mcpolin, Jayant V. Rajan, Colin R. Zamecnik, Ravi Dandekar, Bonny D. Alvarenga, Rita P. Loudermilk, Chloe Gerungan, Collin M. Spencer, Sharon A. Sagan, Danillo G. Augusto, Jessa Alexander, Jill A. Hollenbach, Michael R. Wilson, Scott S. Zamvil, Riley Bove

## Abstract

Vaccine-elicited adaptive immunity is an essential prerequisite for effective prevention and control of coronavirus 19 (COVID-19). Treatment of multiple sclerosis (MS) involves a diverse array of disease-modifying therapies (DMTs) that target antibody and cell-mediated immunity, yet a comprehensive understanding of how MS DMTs impact SARS-CoV-2 vaccine responses is lacking. We completed a detailed analysis of SARS-CoV-2 vaccine-elicited spike antigen-specific IgG and T cell responses in a cohort of healthy controls and MS participants in six different treatment categories. Two specific DMT types, sphingosine-1-phosphate (S1P) receptor modulators and anti-CD20 monoclonal antibodies (mAb), resulted in significantly reduced spike-specific IgG responses. Longer duration of anti-CD20 mAb treatment prior to SARS-CoV-2 vaccination were associated with absent antibody responses. Except for reduced CD4+ T cell responses in S1P-treated patients, spike-specific CD4+ and CD8+ T cell reactivity remained robust across all MS treatment types. These findings have important implications for clinical practice guidelines and vaccination recommendations in MS patients and other immunosuppressed populations.

## Introduction

Multiple sclerosis (MS) is an inflammatory demyelinating condition of the central nervous system (CNS) treated with more than 20 different approved disease modifying therapies (DMTs)^1^. MS DMTs differ considerably in their mechanisms of action with variable impacts on humoral and cellular immune functions leading to associated risks of certain infections^2^. Coronavirus 19 (COVID-19) is an infectious disease caused by severe acute respiratory syndrome coronavirus-2 (SARS-CoV-2), which has resulted in a pandemic since early 2020. Control of SARS-Cov-2 infection involves mobilization of antibody and T cell-mediated immunity^3–5^. Evidence suggests that MS patients on anti-CD20 monoclonal antibody (mAb) therapies are at higher risk for hospitalization from COVID-19^6^. Several recent reports have demonstrated that MS patients treated with anti-CD20 mAb and S1P receptor modulators have reduced or undetectable spike antigen-specific IgG following COVID-19^7–11^.

Vaccines targeting the SARS-CoV-2 spike protein have proven to be highly effective against COVID-19, in which protective immunity involves a combination of robust antibody and CD4+ and CD8+ T cell responses^12–15^. Given the variable effects of different classes of MS DMTs on humoral and cellular immunity, there is a serious concern that SARS-CoV-2 vaccine immunity may be blunted by certain MS treatments and result in increased COVID-19 risk. Indeed, most MS DMTs have been previously reported to at least partially impact vaccine-elicited antibody and/or T cell immunity^16,17^. To date, the majority of studies evaluating SARS-CoV-2 vaccine responses in MS patients have been limited to measuring antibody titers, demonstrating reduced spike antigen-specific antibody responses in MS patients treated with anti-CD20 mAb and S1P receptor modulators^7,18–20^. Several reports have also indicated largely intact spike antigen-specific T cell responses in vaccinated MS patients on anti-CD20 mAb^18,21^. Nevertheless, there is currently no available data comparing SARS-CoV-2 vaccine-specific CD4+ and CD8+ T cell reactivity across patients on different DMTs, presenting a significant gap in our understanding of COVID-19 susceptibility in at-risk patient populations.

The goal of this study was to comprehensively analyze SARS-CoV-2 vaccine-induced humoral and cellular immune responses in MS patients treated with an array of different immunotherapies. Spike antigen-specific IgG and CD4+ and CD8+ T cell responses were measured before and after SARS-CoV-2 vaccination in a cohort of healthy controls (n = 13) and MS patients (n = 67) in six different treatment categories: untreated, glatiramer acetate (GA), dimethyl fumarate (DMF), natalizumab (NTZ), S1P receptor modulators, and anti-CD20 mAb. MS patients treated with anti-CD20 mAb and S1P receptor modulators had substantially reduced levels of total spike IgG and spike receptor-binding domain (RBD)-specific IgG with binding to a restricted array of spike immune determinants. Spike antibody seropositivity in anti-CD20 mAb treated patients was associated with higher CD19+ B cell levels and was inversely correlated with cumulative duration of anti-CD20 mAb therapy. In contrast to antibodies, spike antigen-specific CD4+ and CD8+ T cell responses were overall similar in frequency in all MS treatment groups and with similar cytokine and memory profiles. These findings therefore provide critical insights into the differential effects of MS DMTs on SARS-CoV-2 vaccine-elicited adaptive immunity with important consequences for clinical decision making in vulnerable immunosuppressed patients.

## Materials and Methods

### Study design

In this prospective observational study, participants included patients with clinically definite MS (2017 McDonald Criteria^22^) and healthy controls (not immunocompromised or on immunosuppressive therapy) aged 18-75 years old. All enrolled participants provided written, informed consent for this study, which was approved by the UCSF Committee on Human Research (IRB# 21-33240). MS cohorts included patients not on any treatment (no DMT in the prior 6 months), or treated with GA, DMF, NTZ, any S1P receptor modulator, or intravenous anti-CD20 mAb therapy (RTX or OCR). Only participants with no history of COVID-19 and not previously vaccinated against SARS-CoV-2 prior to enrollment were included. All study participants completed full SARS-CoV-2 vaccination with one of the FDA-approved or authorized vaccines (Comiranty/BNT162b2 from Pfizer/BioNTech, mRNA-1273 from Moderna, or Ad26.COV2 from Johnson and Johnson). Blood samples were collected from all individuals before and two weeks (for Comirnaty/BNT162b2 and mRNA-1273) or four weeks (for Ad26.COV2) following final SARS-CoV-2 vaccination. Basic participant characteristics and vaccine-related variables are outlined in **Table 1**. Treatment-specific characteristics were recorded from the medical record for anti-CD20 mAb and S1P receptor modulator-treated MS patients. For patients on anti-CD20 mAb therapy, total IgG (last measured prior to vaccine), total cumulative treatment duration (i.e. time from start of anti-CD20 treatment until first vaccine dose), and treatment interval between last anti-CD20 mAb infusion and first vaccine dose were recorded. Absolute lymphocyte count (last measured prior to vaccine) and total cumulative treatment duration were also recorded for S1P patients.

**Table 1.**
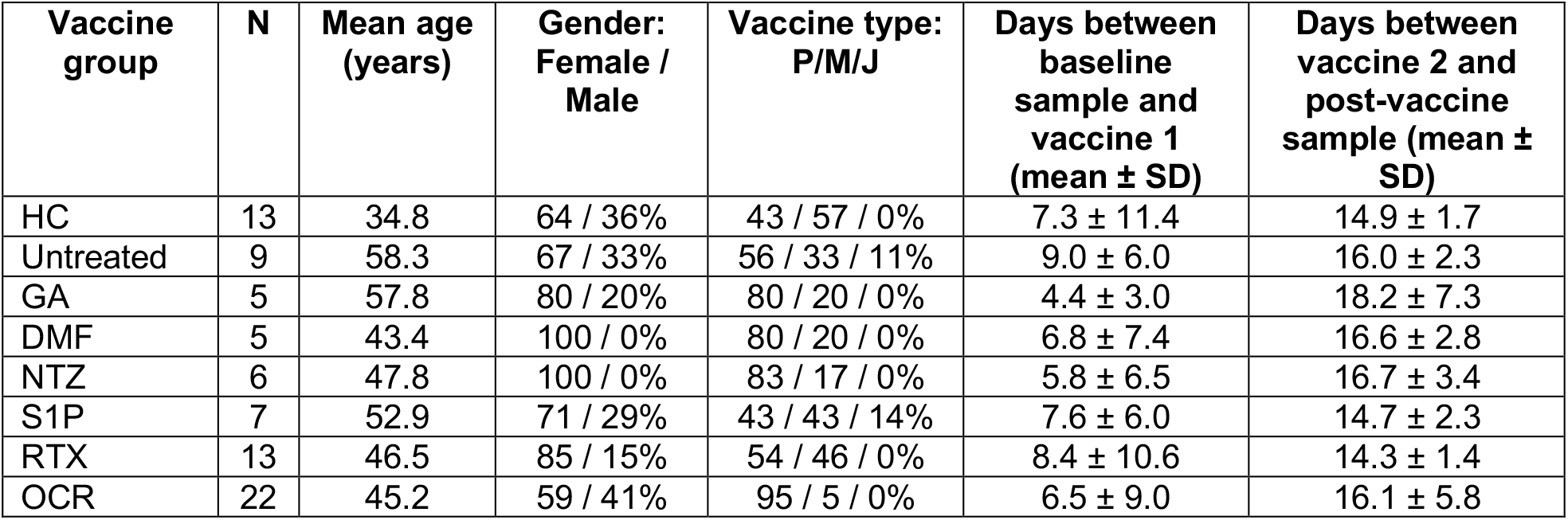
Overview of participants included in the current study. Abbreviations: healthy control (HC), glatiramer acetate (GA), dimethyl fumarate (DMF), natalizumab (NTZ), sphingosine-1-phosphate receptor modulator (S1P), rituximab (RTX), ocrelizumab (OCR), Pfizer/BioNTech (P), Moderna (M), Johnson and Johnson (J).

| Vaccine group | N | Mean age (years) | Gender: Female / Male | Vaccine type: P/M/J | Days between baseline sample and vaccine 1 (mean ± SD) | Days between vaccine 2 and post-vaccine sample (mean ± SD) |
| --- | --- | --- | --- | --- | --- | --- |
| HC | 13 | 34.8 | 64 / 36% | 43 / 57 / 0% | 7.3 ± 11.4 | 14.9 ± 1.7 |
| Untreated | 9 | 58.3 | 67 / 33% | 56 / 33 / 11% | 9.0 ± 6.0 | 16.0 ± 2.3 |
| GA | 5 | 57.8 | 80 / 20% | 80 / 20 / 0% | 4.4 ± 3.0 | 18.2 ± 7.3 |
| DMF | 5 | 43.4 | 100 / 0% | 80 / 20 / 0% | 6.8 ± 7.4 | 16.6 ± 2.8 |
| NTZ | 6 | 47.8 | 100 / 0% | 83 / 17 / 0% | 5.8 ± 6.5 | 16.7 ± 3.4 |
| S1P | 7 | 52.9 | 71 / 29% | 43 / 43 / 14% | 7.6 ± 6.0 | 14.7 ± 2.3 |
| RTX | 13 | 46.5 | 85 / 15% | 54 / 46 / 0% | 8.4 ± 10.6 | 14.3 ± 1.4 |
| OCR | 22 | 45.2 | 59 / 41% | 95 / 5 / 0% | 6.5 ± 9.0 | 16.1 ± 5.8 |

### Sample collection and processing

Blood samples were collected from consented participants at the Neurosciences Clinical Research Unit at UCSF or the patient’s residence through ExamOne (a Quest Diagnostics company). 90 mL of whole blood was collected in heparinized tubes and an additional 10 mL of blood was collected in serum separator tubes at each time point. All samples were processed within 24 hours of collection. Blood samples were spun at 500g for 10 minutes and plasma and serum were removed and frozen at minus 80°C until ready for use. Whole blood pellets were resuspended in 1X DPBS and peripheral blood mononuclear cells (PBMCs) were isolated over Ficoll gradient. PBMCs were frozen in freezing media (10% DMSO and 90% FBS) and stored in liquid nitrogen until the day of experimentation.

### Semi-quantitative spike antibody analysis by Luminex assay

Spectrally distinct Luminex beads were conjugated with trimeric spike protein (residues 1-1213), spike RBD (residues 328-533) (generously provided by Dr. John Pak, Chan Zuckerberg Biohub), or bovine-specific albumin fraction V (BSA) (Sigma-Aldrich #10735094001) at a concentration of 5 µg of protein per 1 million beads. Conjugation was done via an EDC/sulfo-NHS coupling strategy to terminal amines using antibody coupling kit following manufacturer’s instructions (Luminex #40-50016) as performed previously^23^. All serological analyses were performed in duplicate, and beads were pooled on the day of use. Thawed serum samples were diluted in PBS + 0.05% Tween 20 (PBST) containing 1% non-fat milk and mixed with pooled protein-coated beads (2000-2500 beads per ID) at a final serum dilution of 1:500. Samples were incubated for 1 hour at room temperature, washed, and stained with 1:2000 anti-human IgG Fc antibody PE (BioLegend #637310) in PBST for 30 minutes at room temperature. Beads were washed with PBST and analyzed in a 96 well format on a Luminex LX 200 cytometer. The net median fluorescence intensity (net MFI) was recorded for each set of beads. The mean net MFI for total spike and spike RBD for each sample was divided by the net MFI for the corresponding BSA negative control. A net MFI ≥ 5.0 was used as a cutoff for total spike and spike RBD seropositivity, which has been previously demonstrated to be highly sensitive and specific^24^.

### Antibody analysis by coronaphage VirScan

Coronaphage library design and construction, immunoprecipitation and generation of peptide count tables were performed as previously described^23^. All peptide counts were converted to reads/100,000 reads (rp100k). For each vaccinated individual, peptide enrichment was calculated relative to the corresponding pre-vaccination sample as rp100k_post-vaccination_/rp100k_pre-vaccination_. For each sample, enrichments were log transformed, and a Z-score calculated for each peptide in each sample. Peptides with Z-scores greater than 3 in post-vaccination samples were considered significantly enriched over pre-vaccination. Sero-reactivity maps were generated for each sample by aligning each significantly enriched peptide to the concatenated open reading frames of SARS-CoV-2, focusing on the spike protein. Signal intensity at each position in the spike protein was the sum of signal for all peptides covering each position and was used to generate heatmaps as well as plots depicting the proportion of individuals with sero-reactivity at each position in each treatment group.

### Flow cytometry analysis of basic immune cell subsets

PBMCs were thawed, washed, and equilibrated in RPMI with 10% FBS for 2 hours at 37°C and stained with the indicated cell surface panel for identifying immune cell subsets as shown in **Supplemental Table 1**. All samples were collected on an LSR Fortessa (BD). The gating strategy employed is shown **Supplemental Figure 1A**. Flow cytometry analysis was completed using FlowJo (BD).

### T cell analysis by activation-induced marker (AIM) expression and intracellular cytokine stimulation (ICS)

PBMCs were thawed, washed, and equilibrated in RPMI with 10% FBS for 2 hours at 37°C prior to initiation of functional T cell studies. PBMCs were washed and resuspended in serum-free RPMI for AIM studies or resuspended in serum-free RPMI containing 1:500 Golgi-Stop (BD), 1:500 Golgi-Plug (BD), and 1:200 Fast-immune (BD) for ICS studies. For all studies, PBMCs were plated at 1 × 10^6^ cells per well in 96-well round-bottom plates. PBMCs were stimulated in parallel with spike peptide pools (2 pools, 157 and 158 peptides, JPT Peptide Technologies) at a final concentration of 1 µg/ml per peptide. 0.2% DMSO vehicle control was used for no stimulation in all assays. PBMCs were stimulated for 24 hours for AIM assays and 16 hours for ICS assays. Cells were washed with FACS wash buffer (1X DPBS without calcium or magnesium, 0.1% sodium azide, 2 mM EDTA, 1% FBS) and stained with the indicated antibody panels for AIM and ICS in **Supplemental Table 1**. In the case of AIM assays, cells were washed with FACS wash buffer, fixed with 2% paraformaldehyde (BD), and stored in FACS wash buffer in the dark at 4°C until ready for flow cytometry analysis as above. For ICS assays, cells were washed after cell surface staining and stained with a cocktail of intracellular cytokine antibodies (see **Supplemental Table 1** for antibody panel) in permeabilization buffer for 1-2 hours at 4°C. ICS samples were then fixed, washed, and stored as done for AIM samples until ready for flow cytometry analysis. The gating strategy for AIM and ICS is show in **Supplemental Figures 3A-B**. The frequencies of spike-specific T cells were calculated by subtracting the no stimulation background from the corresponding S1 and S2 pool-stimulated samples, which were then summed together.

### HLA genotyping

Genomic DNA was isolated using the QiaAmp DNA Blood Mini kit (Qiagen). A total of 100 ng of high-quality DNA was fragmented using the KAPA HyperPlus kit (Roche). Subsequently, the fragmented DNA had their ends repaired, poly-A tail added and ligated through PCR to Illumina-compatible dual index adapters uniquely barcoded. After ligation, fragments were purified with 0.8X ratio AMPure XP magnetic beads followed by double size selection (0.42X and 0.15X ratios) to select libraries of approximately 800 bp. Finally, libraries were amplified and purified with magnetic beads. After fluorometric quantification, 30 ng of each sample were precisely pooled using ultrasonic acoustic energy, and the targeted capture was performed with HyperCap kit (Roche). Briefly, the volumes were reduced using magnetic beads, and the DNA libraries were bound to 1,394 biotinylated probes specific to the *HLA* region, covering all exons, introns, and regulatory regions of *HLA-A, HLA-B, HLA-C, HLA-DRB1, HLA-DRA, HLA-DQB1, HLA-DQA1, HLA-DPB1*, and *HLA-DPA1*. Fragments targeted by the probes were captured with streptavidin magnetic beads and then amplified and purified. Enriched libraries were analyzed in BioAnalyzer (Agilent) and quantified by digital-droplet PCR. Finally, enriched libraries were sequenced with the HiSeq4000 platform (Illumina) with paired-end 150 bp sequencing protocol. After sequencing, data were analyzed with the software HLA Explorer (Omixon).

### Spike antigen-specific CD8+ T cell analysis by pMHC tetramer

pMHC I tetramers loaded with spike peptides and labeled with the fluorophores listed in **Supplemental Table 2** were generated from UV-photolabile monomers for HLA-A*01:01, HLA-A*02:01, HLA-A*03:01, HLA-A*11:01, and HLA-B*07:02 monomers (NIH Tetramer Core) by UV peptide exchange as previously described^25,26^. 500 µM D-biotin was added to each tetramer and tetramers were pooled as indicated in **Supplemental Table 2** on the day of use. All tetramer experiments were completed within three weeks of tetramer generation. For each tested sample, 2-3 × 10^7^ PBMCs were thawed, washed and equilibrated in RPMI with 10% FBS for 1 hour at 37 °C. The frequencies of spike antigen-specific CD8^+^ T cells were calculated as previously described^26,27^. In brief, an aliquot of PBMCs was used for cell surface staining (panel listed in **Supplemental Table 1**) and counted with 123count eBeads (Invitrogen) prior to tetramer enrichment. The remainder of PBMCs were stained with the indicated tetramer pools for 30 minutes at room temperature, washed, and enriched using anti-PE magnetic microbeads (Miltenyi) over a magnetic column. Tetramer-enriched cells were cell surface-stained and counted as done for pre-enrichment. The gating strategy is outlined in **Supplemental Figure 3C**; a stringent tetramer gating strategy was employed whereby CD8+ T cells labeled with only two fluorophores were considered antigen-specific (i.e. cells that stained positive with more or less than two fluorophores were excluded from the analysis). Spike tetramer-positive CD8+ T cells with frequencies greater than 1 × 10^−5^ per total CD8+ T cells were considered positive.

### Statistical analysis

Pre-vaccine and post-vaccine antibody and T cell responses were compared by multiple paired two-way t-tests. Univariate analysis by serostatus was performed by Mann-Whitney test. 2-way ANOVA with Tukey multiple comparisons was used to analyze post-vaccination antibody and T cell responses across different groups; untreated MS patients were used as the comparison group for statistical significance unless stated otherwise. Simple linear regression was to analyze IgG levels with the indicated independent variables and Spearman rank was used for correlation analysis. The following *p*-values were used: not significant (ns) for *p* ≥ 0.05, * for *p* < 0.05, ** for *p* < 0.01, *** for *p* < 0.001, **** for *p* < 0.0001.

## Results

### Study overview

To study the effects of different MS DMTs on SARS-CoV-2 vaccine-induced immune responses, we recruited a cohort of 80 participants comprised of healthy controls (HC, n = 13) and MS patients on no treatment (n = 9) or treated with GA (n = 5), DMF (n = 5), NTZ (n = 6), S1P (n = 7), or anti-CD20 mAb, including rituximab (RTX, n = 13), or ocrelizumab (OCR, n = 22) (**Table 1**). Baseline samples were collected prior to SARS-CoV-2 vaccination (mean 7.2 days, range 0-34 days before first vaccine) and post-vaccine samples were collected approximately two weeks following the second mRNA COVID-19 vaccine (BNT162b2 or mRNA-1273) (mean 15.7 days, range 11-40 days) or four weeks following adenoviral vaccine (Ad26.COV2.S) (mean 28.5 days, range 28-29 days).

### Overview of basic immune cell subsets

The percentages of immune cell subsets, including CD4+ and CD8+ T cells, CD19+ B cells, CD14+, CD14+ CD16+, and CD16+ cells, were evaluated in all participants before and after SARS-CoV-2 vaccination using the gating strategy shown in **Supplemental Figure 1A**. Except for CD14+ CD16+ cells in GA-treated patients (*p* = 0.0425), no significant differences were observed in any of the immune cell subsets before and after SARS-CoV-2 vaccination in all other cohorts (**Supplemental Figure 1B-G**). While the percentage of CD8+ T cells were not significantly affected by treatment status, the percentage of CD4+ T cells were significantly reduced before (*p* < 0.0001) and after (*p* < 0.0001) vaccination in S1P-treated compared to untreated MS patients, consistent with the known mechanism of action of S1P receptor modulators^28^ (**Supplemental Figure 1B**). As expected, the percentages of CD19+ B cells were also significantly reduced at both collection time points in S1P (*p* < 0.0001), RTX (*p* < 0.0001), and OCR (*p* < 0.0001) patients compared with untreated MS patients (**Supplemental Figure 1D**).

### Semi-quantitative analysis of anti-spike antibodies by MS treatment type

Total spike IgG and spike RBD IgG were measured as net MFI after normalization to background BSA control^23^. Healthy controls, untreated MS, and MS patients treated with GA, DMF, and NTZ demonstrated significantly increased total spike IgG (**Figure 1A**) and spike RBD IgG (**Figure 1B**) levels following SARS-CoV-2 vaccination compared to their respective pre-vaccination time points. Of note, the one untreated MS patient who received the single injection Ad26.COV2.S vaccine also had the lowest total spike and spike RBD IgG antibody. In contrast, vaccine-elicited total spike and spike RBD IgG levels were variable amongst S1P, RTX, and OCR patients, with undetectable antibody levels in some patients and near normal IgG levels in others (**Figure 1A-B**). Overall, S1P and RTX patients showed no significant increase in post-vaccination total spike IgG compared to pre-vaccination whereas spike RBD IgG in S1P, RTX, and OCR were not significantly increased following COVID-19 vaccination (**Figure 1A-B**). Post-vaccination total spike IgG levels in HC and MS patients treated with GA, DMF, NTZ were similar to untreated MS, but spike RBD IgG was significantly higher in DMF (*p* = 0.038) and NTZ (*p* < 0.0001). In contrast, post-vaccination total spike and spike RBD IgG were significantly reduced in MS patients treated with S1P, RTX, and OCR (**Figure 1A-B**). IgG seropositivity to total spike protein and spike RBD following SARS-CoV-2 vaccination were also compared between untreated MS patients and all other cohorts. Only RTX patients had a significant decrease in total spike IgG seropositivity (23.1% ± 12.2% SEM, *p* < 0.0001), whereas S1P, RTX, and OCR patients had significant reductions in spike RBD IgG seropositivity (42.9 ± 20.2% SEM, *p* = 0.02; 7.7 ± 7.7% SEM, *p* < 0.0001; 36.4 ± 10.5% SEM, *p* = 0.0001, respectively) (**Figure 1C**). These findings are consistent with recent reports indicating reductions in SARS-CoV-2 spike-specific antibodies in SARS-CoV-2 vaccinated MS patients on S1P receptor modulators and anti-CD20 mAb^7,18,19^.

**Figure 1.**
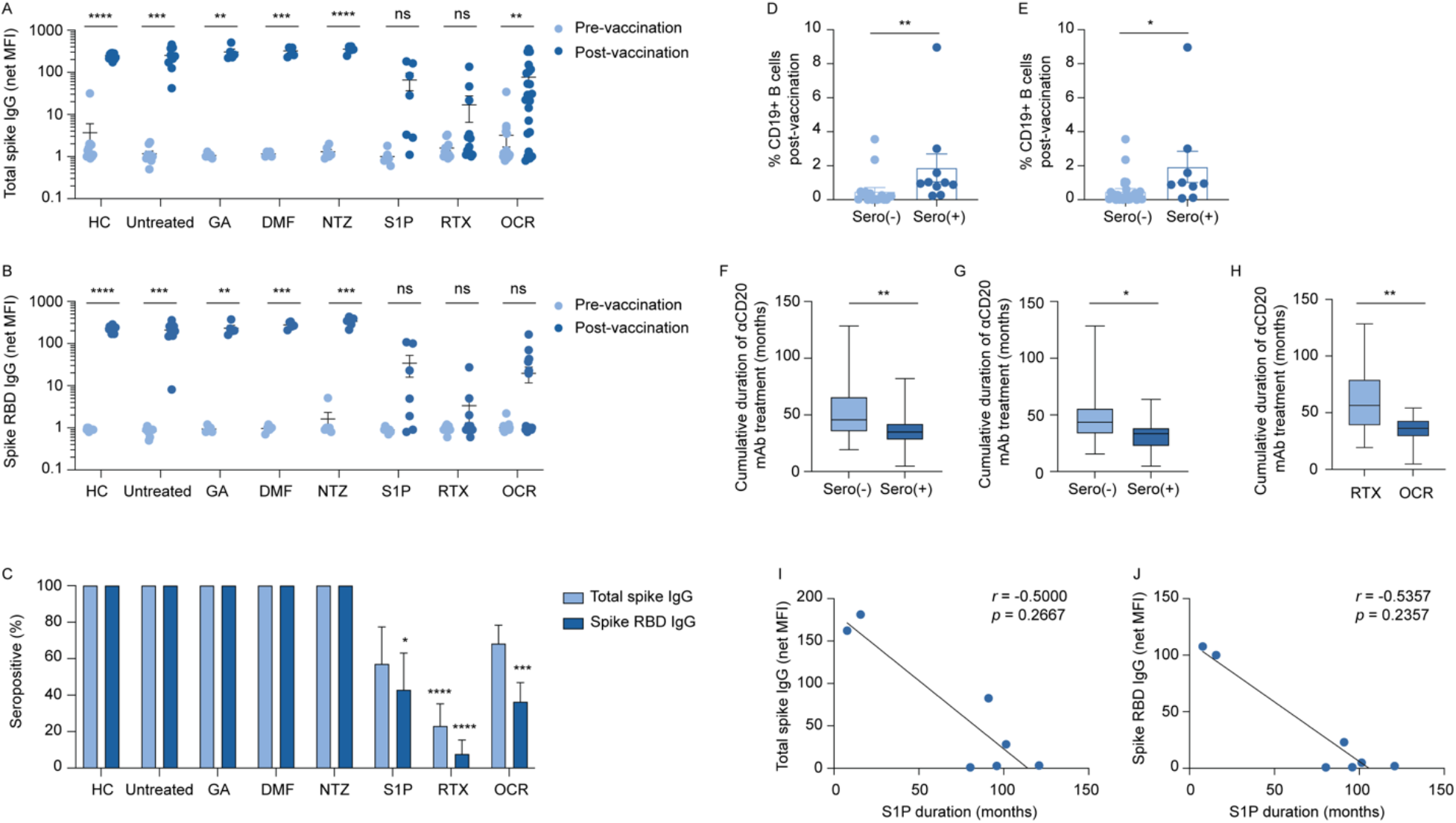
Analysis of total spike and spike RBD IgG before and after SARS-CoV-2 vaccination of MS patients on different DMTs. Net MFI of total spike IgG (**A**) and spike RBD IgG (**B**) at pre- and post-vaccination time points (multiple paired t-tests). Percent seropositivity of total spike IgG and spike RBD IgG following vaccination for each cohort (**C**) (2-way ANOVA with Tukey multiple comparisons; significance based on comparison between untreated MS and other MS treatment groups). Percent CD19+ B cells following vaccination in total spike IgG (**D**) and spike RBD (**E**) seronegative and seropositive anti-CD20 mAb patients (Mann-Whitney). Cumulative duration of anti-CD20 mAb treatment prior to SARS-CoV-2 vaccination by serologic status of total spike IgG (**F**) and spike RBD (**G**) and by anti-CD20 mAb type (**H**) (Mann-Whitney). Simple linear regression of net MFI of total spike IgG (**I**) and spike RBD IgG (**J**) by duration of S1P receptor modulator duration (correlation by Spearman rank).

### Factors associated with anti-spike IgG seropositivity in anti-CD20 mAb and S1P receptor modulator-treated MS patients

Given the variations in total spike and spike RBD IgG levels in MS patients on anti-CD20 mAb, we investigated factors associated with SARS-CoV-2 vaccine-elicited antibody responses. Within the anti-CD20 mAb cohorts (RTX and OCR), we employed a univariate analysis of anti-spike antibody responses by gender, vaccine type, cumulative treatment duration prior to vaccination, total IgG levels, interval between last anti-CD20 infusion and first vaccine dose, lymphocyte subsets before and after vaccination, and HLA-DRB1*15:01 status, given its well established association with MS susceptibility^29^. There was no association with gender, mRNA-vaccine type, or last measured total IgG levels with total spike or spike RBD IgG levels (**Supplemental Figure 2A-F**). There was also no relationship between HLA-DRB1*15:01 status or pre- and post-vaccination percentages of CD4+ and CD8+ T cells and total spike IgG or spike RBD IgG. While there was not a significant difference between the percentage of pre-vaccination CD19+ B cells and total spike and spike RBD serostatus, there was a significant increase in post-vaccination CD19+ B cells and total spike (*p* = 0.0021) and spike RBD (*p* = 0.0158) seropositive patients compared to those who were seronegative (**Figure 1D-E**). Similarly, although there was no significant correlation with total spike and spike RBD IgG and percentages of CD19+ B cells prior to vaccination (**Supplemental Figure 2G-H**), there was a strong positive correlation between the percentage of post-vaccination CD19+ B cells and total spike IgG (*r* = 0.6084, *p* = 0.0001) and spike RBD IgG (*r* = 0.4166, *p* = 0.0128) (**Supplemental Figure 2I-J**). The mean interval between blood sample collection at pre-vaccination and post-vaccination in anti-CD20 mAb patients was 6.6 weeks, therefore we reasoned a potential difference for the discrepancy in correlations of CD19+ B cell levels at the two time points was possible interim B cell reconstitution during the vaccination period. We calculated the differences in percentages of CD19+ B cells between the post-vaccine and pre-vaccine time points and observed a significant correlation between CD19+ B cell changes and total spike (*r* = 0.4903, *p* = 0.0028) and spike RBD (*r* = 0.4005, *p* = 0.0171) IgG levels (**Supplemental Figure 2K-L**), providing support for this hypothesis.

Given variability in timing of anti-CD20 mAb infusions caused by the ongoing SARS-CoV-2 pandemic, we also assessed whether differences in the interval between the last prior anti-CD20 mAb infusion and first SARS-CoV-2 vaccination could be related to vaccine-induced IgG responses, which has been reported previously^18,19^. In the combined anti-CD20 mAb cohorts, we did not observe any significant correlation between infusion to vaccination interval and total spike and spike RBD IgG levels (**Supplemental Figure 2M-N**). We also assessed the cumulative duration of anti-CD20 mAb treatment (i.e. time from start of anti-CD20 mAb treatment until first SARS-CoV-2 vaccination). We found that total spike and spike RBD seronegative patients had a significantly longer cumulative mAb treatment duration (median 45.7 months and 43.5 months, respectively) compared to seropositive patients (median 34.9 months and 33.4 months, respectively) (**Figure 1F-G**). Notably, the significant reduction in total spike IgG seropositivity in RTX compared to OCR patients (**Figure 1C**) was explained by a significantly increased cumulative duration of therapy in the RTX compared to OCR cohorts (median 56.4 months and 36.4 months, respectively; *p* = 0.0017) (**Figure 1H**).

We also assessed factors contributing to variability in anti-spike seropositivity in patients on S1P receptor modulators. Although a smaller cohort, we observed a non-significant trend between longer cumulative duration of S1P receptor modulator therapy and lower total spike and spike RBD IgG (**Figure 1I-J**). There was no significant relationship between total spike and spike RBD IgG and absolute lymphocyte count, CD19+ B cells, CD4+ T cells, or CD8+ T cells.

### Identification of anti-spike protein IgG determinants by coronavirus VirScan

We sought to further explore the immune determinants of SARS-CoV-2 vaccine-induced IgG using VirScan analysis of patient sera from pre- and post-vaccination time points. Sera was probed against a library of overlapping peptides (38 amino acids each) spanning the entire proteomes of 9 different human coronaviruses (including SARS-CoV-2) as well as a peptide library of the spike protein of SARS-CoV-1 and SARS-CoV-2, as previously demonstrated in COVID-19 patients^23^. No signal was detected against spike peptide sequences from any of the pre-vaccinated sera, indicating that all measured spike-specific IgG responses resulted from SARS-CoV-2 vaccination rather than pre-existing cross-reactive immunity against other coronaviruses. Multiple antibody binding determinants were revealed throughout the spike protein, including against the N-terminal domain (NTD), regions flanking the RBD, the S1/S2 cleavage site, fusion site, as well as C-terminal region, in healthy controls and untreated MS patients (**Figure 2**). Of note, neutralizing antibodies targeting RBD epitopes are largely conformation-dependent^30^, which is generally not well represented by phage-displayed peptide due to use of linear peptide sequences. Anti-spike antibody reactivity was slightly more narrowed in GA, DMF, and NTZ patients, with some loss of reactivity in subdominant NTD and C-terminal regions. In anti-CD20 mAb and S1P patients, however, sero-reactivity against the spike protein was far more restricted, primarily against determinants flanking the RBD and the C-terminal region (**Figure 2**). These findings therefore highlight that anti-CD20 mAb and S1P receptor modulator treatments may lead to qualitative changes in anti-spike IgG in addition to quantitative changes.

**Figure 2.**
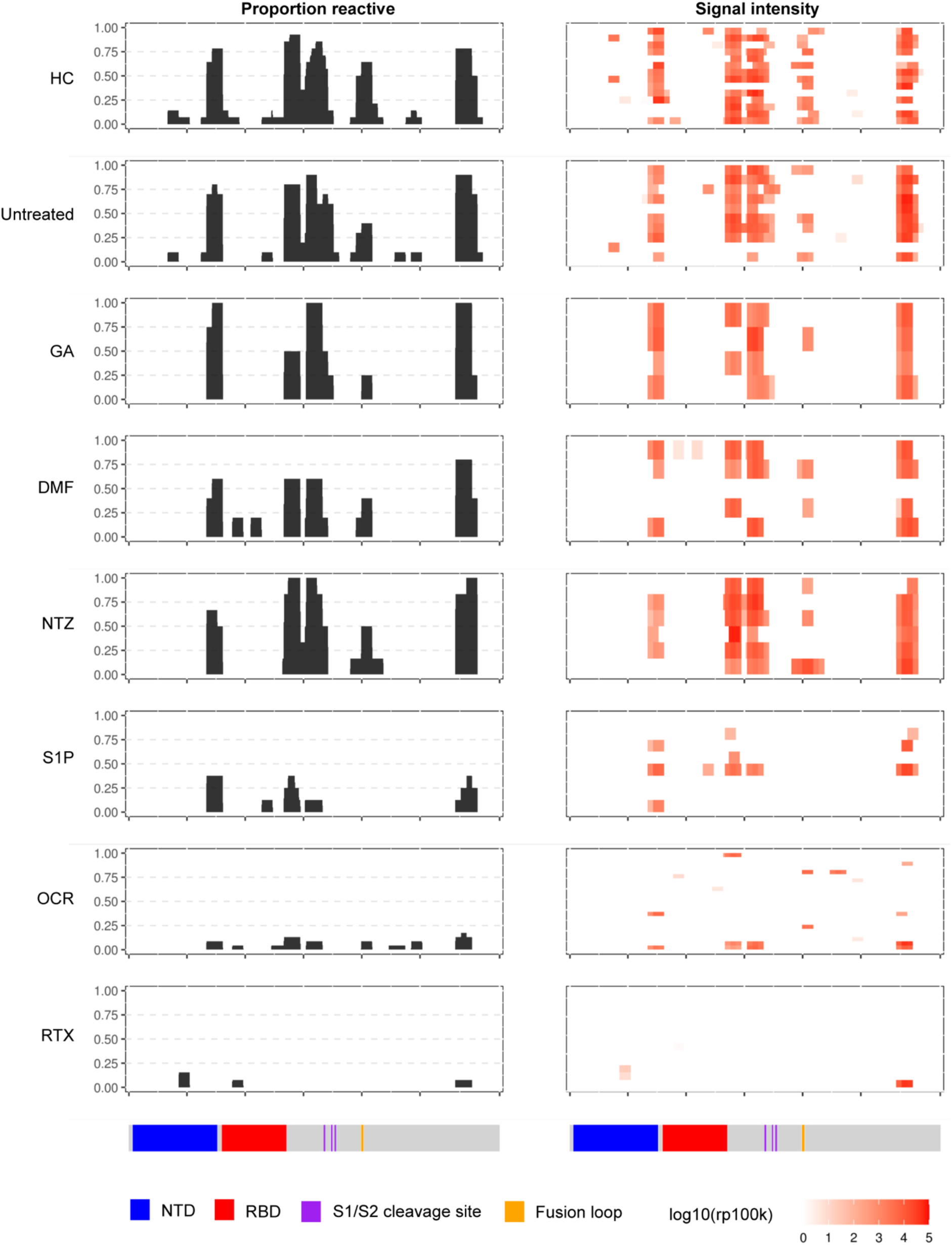
VirScan analysis of post-vaccination antibody reactivity against the SARS spike proteome by MS DMT status. The left column indicates the proportion of individuals sero-reactive against the different regions of the spike protein and the right column indicates the relative signal intensity of antibody binding where each individual is separated by row. The corresponding regions of the spike protein are indicated below the plots.

### Evaluation of spike antigen-specific CD4+ and CD8+ T cells

We investigated the frequency and phenotype of spike antigen-specific CD4+ and CD8+ T cells using pools of spike peptides spanning the entire spike protein by activation-induced marker (AIM) expression and intracellular cytokine stimulation (ICS) (gating strategies shown in **Supplemental Figure 3**). In the AIM assay, antigen reactivity was assessed by CD137 and OX-40 co-expression in CD4+ T cells and CD137 and CD69 co-expression by CD8+ T cells (**Figure 3A and 3D**), as previously demonstrated^31^. Cytokine analysis included IFNγ, TNFα, and IL-2, the dominant cytokines produced by spike-specific T cells^31,32^, as well as IL-4 and IL-10, which can be upregulated by certain MS DMTs^33^. A significant increase in spike-specific CD4+ T cells was observed by AIM in all post-vaccination groups apart from the S1P cohort (**Figure 3B**), likely due to the pronounced S1P-mediated CD4+ T cell lymphopenia. Importantly, none of the post-vaccination MS treatment groups showed a significant reduction in spike-specific CD4+ T cells compared to untreated MS patients (**Figure 3B**). CD4+ T cells from all post-vaccination cohorts produced similar frequencies of IFNγ, TNFα, and IL-2, indicating broad polyfunctionality regardless of MS treatment status (**Figure 3C**). In contrast, frequencies of IL-4- and IL-10-producing CD4+ T cells were minimal with no changes in any of the DMT MS cohorts. Spike antigen-specific CD8+ T cells were significantly increased in all post-vaccination MS cohorts except for GA-treated patients (**Figure 3E**), which was likely influenced by the higher variability and smaller sample size in that group. Moreover, none of the post-vaccination MS treatment groups showed a significant reduction of spike-specific CD8+ T cells measured by AIM compared to untreated MS patients (**Figure 3E**). Cytokine production by post-vaccination spike antigen-specific CD8+ T cells revealed similar polyfunctionality with significant production of IFNγ, TNFα, and IL-2, but minimal IL-4 and IL-10 production (**Figure 3F**). While cytokine responses were overall similar between all MS treatment cohorts, a significant increase in IFNγ+ CD8+ T cells was observed in RTX-treated patients compared to untreated MS patients (**Figure 3F**). In addition, no significant relationship was found between the frequencies of spike-specific CD4+ and CD8+ T cells measured by AIM and total spike and spike RBD IgG seropositivity in anti-CD20 mAb-treated patients.

**Figure 3.**
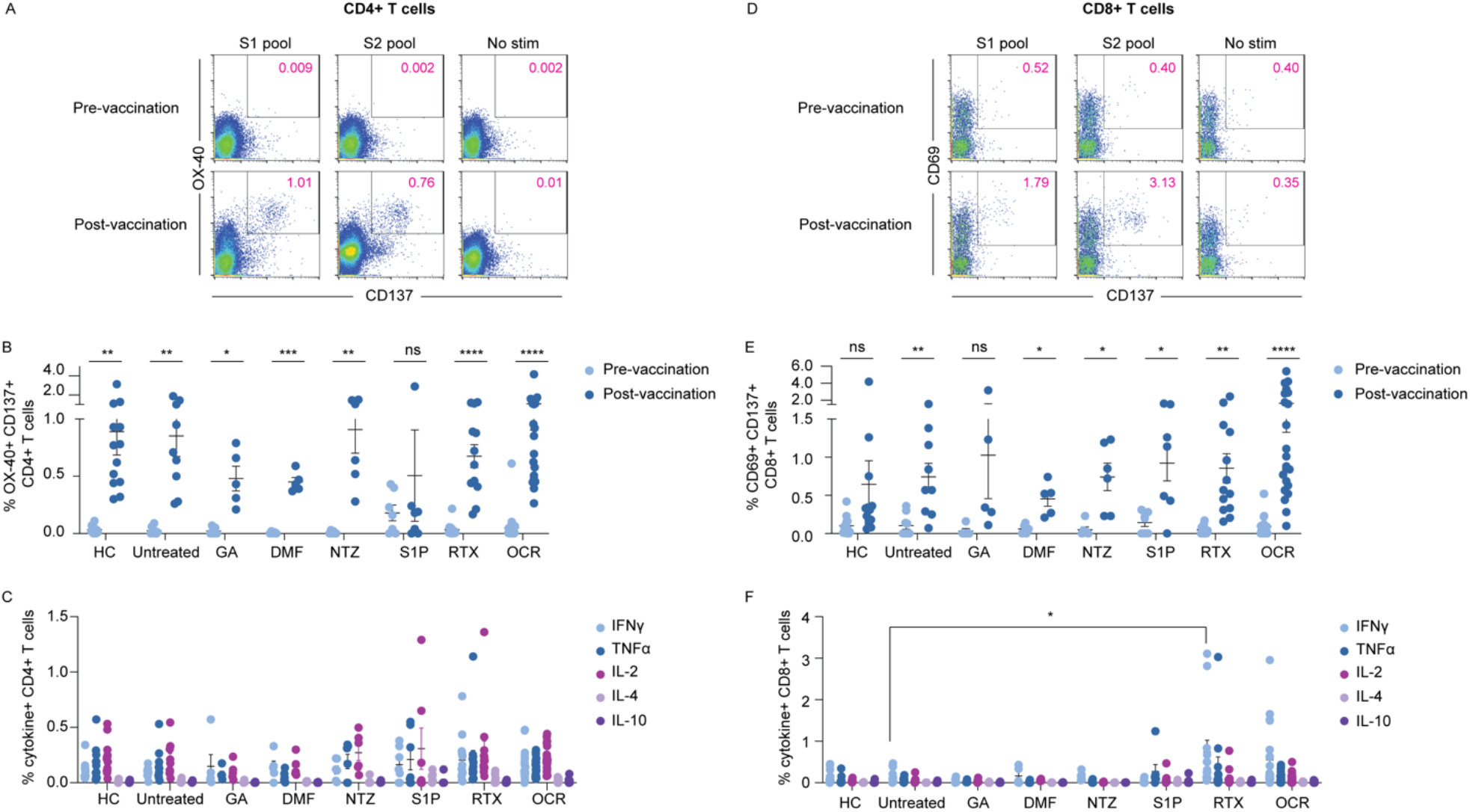
Evaluation of spike antigen-specific CD4+ and CD8+ T cells in MS patients on different DMTs. AIM analysis of CD4+ (**A**) and CD8+ (**D**) T cells from a representative MS patient before and after SARS-CoV-2 vaccination. Summarized AIM and ICS analysis of CD4+ (**B, C**) and CD8+ T cells (**E, F**) across all cohorts. AIM data are shown for pre- and post-vaccination time points (multiple paired t-tests); ICS data depicts post-vaccination analysis only (two-way ANOVA with Tukey multiple comparisons).

### Ex vivo evaluation of spike antigen-specific CD8+ T cells by pMHC tetramers

To further characterize the individual SARS-CoV-2 vaccine-elicited CD8+ T cell response at the individual epitope level, *ex vivo* analysis of spike-specific CD8+ T cells by pMHC tetramers was performed in a subset of post-vaccination participants from HC and each MS cohort (**Supplemental Table 3**). Several panels of pMHC tetramers were generated using previously published spike epitopes^15,32,34–36^. Combinatorial tetramer staining (**Supplemental Table 2**) and enrichment were performed as previously described^26,37^. Spike-specific CD8+ T cells identified by tetramer enrichment were subsequently assessed for memory status by CCR7 and CD45RA expression (**Figure 4A**). The proportion of samples with detectable spike tetramer-positive CD8+T cells was similar across all MS cohorts, ranging from 27-56% tetramer positivity (**Figure 4B**). The mean frequencies of spike tetramer-positive CD8+ T cells did not significantly differ between MS cohorts (**Figure 4C**), although there were variations in spike-specific CD8+ T cell population sizes, which was at least partially related to differences in the *HLA* genotypes available for tetramer analysis across the different patient cohorts (**Supplemental Table 3**). Consistent with a post-vaccination response measured in the peripheral blood, spike-specific CD8+ T cells were predominantly effector memory (T_EM_), which were significantly higher than corresponding naïve T cell (T_N_) populations in all cohorts (**Figure 4D**). In addition, the proportion of T_N_, T_CM_, T_EM_, and T_EMRA_ spike-specific CD8+ T cells did not significantly differ across any of the MS cohorts. Overall, these findings indicate that the magnitude and activation state of SARS-CoV-2 vaccine-elicited T cell responses are not substantially changed by the various MS immunotherapies evaluated in this study.

**Figure 4.**
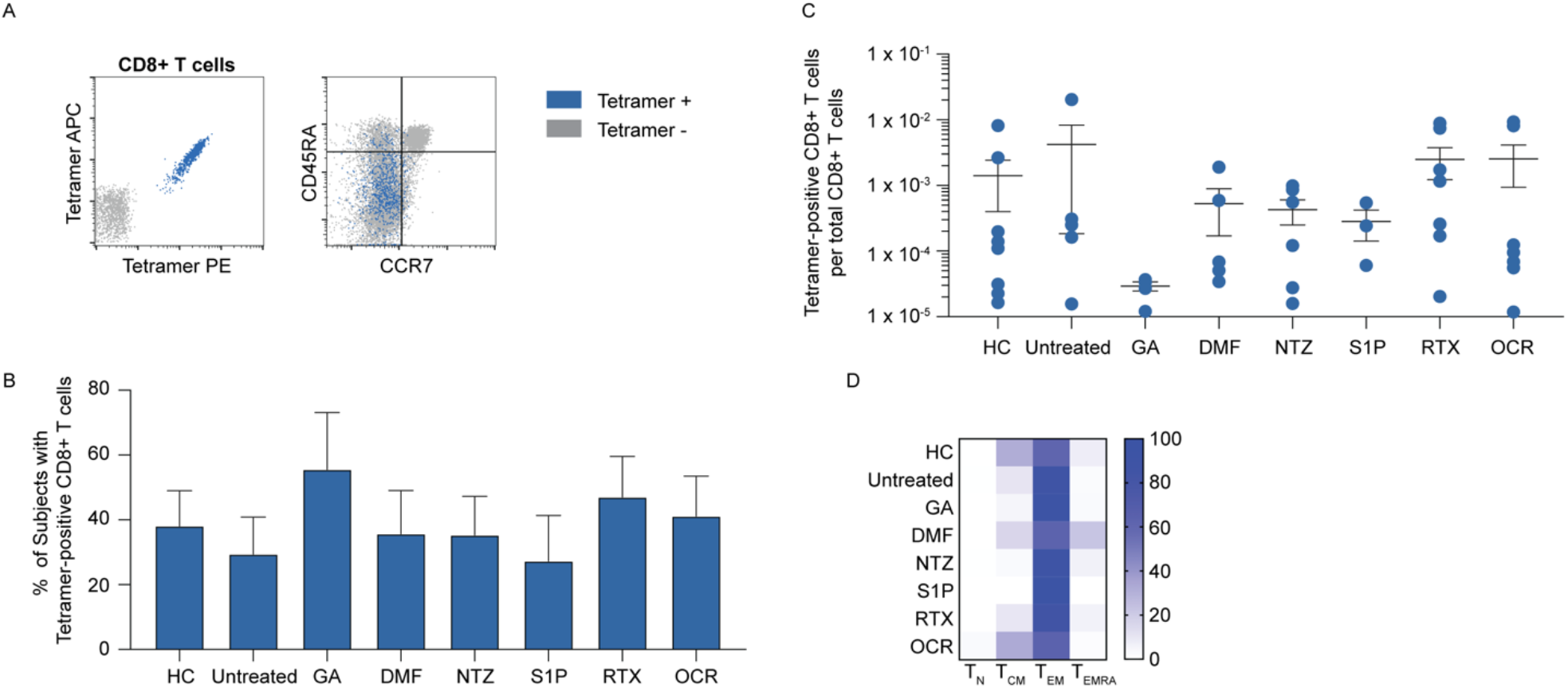
*Ex vivo* analysis of post-vaccination spike-specific CD8+ T cells of MS patients on different DMTs by pMHC I tetramer. Representative analysis of enriched spike peptide tetramer-positive CD8+ T cells (left panel) and memory analysis by tetramer status (right panel) (**A**). The proportion of tested subjects in each cohort with detectable spike tetramer-positive CD8+ T cells (**B**) and their frequencies (**C**) are shown. Heat-map analysis of memory subsets of spike tetramer-positive CD8+ T cells in each cohort (**D**).

## Discussion

MS DMTs differentially impact humoral and cell-mediated immunity, both of which are essential for protection against COVID-19^3,5^. In support of this, recent data suggests that unvaccinated MS patients on anti-CD20 mAb treatments are at higher risk for severe COVID-19^6^. To date, the majority of studies evaluating SARS-CoV-2 vaccine responses in MS patients have been limited to measuring antibody levels^8–11^ and those that have explored T cell reactivity have primarily focused on anti-CD20 mAb-treated patients^18,21^. Thus, there is a significant need to comprehensively investigate how different MS DMTs impact SARS-CoV-2 vaccine-elicited antibody and CD4+ and CD8+ T cell immunity.

Following vaccination, untreated MS patients and those treated with GA, DMF, and NTZ mounted similar total spike and spike RBD IgG responses compared to healthy controls. In contrast, patients on S1P receptor modulators and anti-CD20 mAb had significantly reduced levels of total spike and spike RBD IgG, consistent with recent reports^8–11^. We did not observe a clear relationship between spike antibody seropositivity and anti-CD20 mAb infusion interval and SARS-CoV-2 vaccination, in contrast to several other studies^7,18,19,21^. It is possible this disparity could be due to differences in patient populations or methodology. Our finding that the percentage of CD19+ B cells following vaccination, but not prior to vaccination, was significantly associated with spike antibody seropositivity suggests that interval B cell reconstitution occurring in lymphoid tissue prior to circulation in the blood may be sufficient for antibody generation. Indeed this is supported by a recent finding that spike IgG seropositivity is present in a portion of anti-CD20 mAb-treated MS patients despite the absence of circulating spike-specific B cells^18^. Furthermore, the negative effect of cumulative anti-CD20 mAb treatment duration on spike IgG seropositivity suggests that while essentially all circulating B cells are rapidly depleted following anti-CD20 mAb treatment, B cells may initially persist in smaller numbers in secondary lymphoid tissue^38^ but are ultimately depleted with long-term treatment.

A novel strength of our study was the ability to assess high resolution antibody reactivity across the entire spike protein using programmable phage display (VirScan). Antibody reactivity against a broad range of spike determinants was observed amongst healthy controls and untreated MS patients, including regions vital for SARS-CoV-2 entry into cells. In contrast, antibody reactivity was restricted to a narrower range of spike determinants in seropositive S1P and anti-CD20 mAb-treated MS patients. These data therefore suggest that there are qualitative differences in SARS-CoV-2 vaccine-elicited antibody responses for certain MS DMTs with potential important implications on antibody functional efficacy in patients with blunted spike sero-reactivity.

We also performed an extensive analysis of spike-specific CD4+ and CD8+ T cells in all participants before and after SARS-CoV-2 vaccination. In contrast to the humoral response, spike-specific CD4+ and CD8+ T cell responses were largely intact across all MS cohorts irrespective of DMT status. Vaccine-elicited CD4+ T cell responses were diminished in S1P patients, consistent with the preferential reduction of circulating CD4+ T cells by S1P receptor modulators^28^. In addition, spike-specific CD4+ and CD8+ T cells from all MS treatment groups produced multiple effector cytokines, suggesting DMT exposure did not alter T cell polyfunctionality. Interestingly, we observed a trend towards increased CD8+ T cell cytokine production in anti-CD20 mAb-treated compared to untreated MS patients, which was significantly increased in the case of IFNγ+ CD8+ T cells in the RTX cohort. These findings support recent similar reports of increased SARS-CoV-2-specific CD8+ T cell activation in anti-CD20 mAb-treated MS patients^18,21^ and suggest that B cell depletion may result in compensatory changes in certain aspects of cellular immunity. However, the mechanism of such T cell-mediated changes and whether this has a protective effect against COVID-19 remain unknown.

The results of our study have significant potential implications for clinical guidance in MS patients and other vulnerable immunocompromised patient populations. The finding that certain MS immunotherapies preferentially disrupt humoral immune responses following SARS-CoV-2 vaccination raises the concern that patients on such therapies may be at higher risk of COVID-19^39^. In addition, our data highlight the cumulative negative impact of prolonged anti-CD20 mAb treatment on the generation of *de novo* humoral immunity. On the other hand, the preservation of cell-mediated immunity provides reassurance that all immunosuppressed MS patients will obtain at least partial protection from more severe COVID-19 outcomes. An outstanding question is whether immunosuppressed MS patients will benefit from booster SARS-CoV-2 vaccinations, either by antibody seroconversion and/or augmentation of cell-mediated immunity. The data from our study therefore provide important insights regarding COVID-19 risk assessment and SARS-CoV-2 vaccination practices for immunosuppressed patient populations.

## Data Availability

All data are available in the main text or the supplementary materials. Raw data are available from the authors upon request.

## Acknowledgments

We thank all study participants for their generosity and willingness to contribute to this study. This work was supported by grants from the NIH 1K08NS107619 (JJS), R01NS092835 (SSZ), R01AI131624 (SSZ), R21NS108159 (SSZ), NMSS TA-1903-33713 (JJS), RG1701-26628 (SSZ), and the Maisin Foundation (SSZ).

## Author Contributions

JJS, RB, SSZ, and MRW designed and supervised the study. WR, K Mcpolin, and JA performed clinical recruitment and sample acquisition. Patient data was collected and managed by JJS, WR, and K Mcpolin. JJS, K Mittl, WR, CG, CMS, and SAS carried out sample processing. Luminex analysis was completed by JJS, K Mittl, CRZ, RL, and CG. BA completed VirScan library preparation and JVR and RD performed VirScan analysis. JJS and K Mittl performed all T cell studies. DGA and JAH performed HLA genotyping. Statistical analysis was completed by JJS and JAH. JJS wrote the initial draft of the manuscript. All authors contributed to data interpretation and manuscript review.

