## Supplemental Data for "Impact of multiple sclerosis disease-modifying therapies on SARS-CoV-2 vaccine-induced antibody and T cell immunity"

| Immune cell subset panel | | | | |
|---|---|---|---|---|
| **Antibody target** | **Clone** | **Fluorophore** | **Vendor** | **Catalog #** |
| CD3 | OKT3 | Alexa 647 | BioLegend | 317312 |
| CD4 | A161A1 | PE | BioLegend | 357404 |
| CD8 | SK1 | Alexa 700 | BioLegend | 344724 |
| CD14 | HCD14 | BV421 | BioLegend | 325628 |
| CD16 | B73.1 | PE-Cy7 | BioLegend | 360708 |
| CD19 | HIB19 | PerCP-Cy5.5 | BioLegend | 302230 |
| Live/dead dye | | eFluor 506 | Invitrogen | 65-0866-14 |
| **Activation induced marker (AIM) panel** | | | | |
| **Antibody target** | **Clone** | **Fluorophore** | **Vendor** | **Catalog #** |
| CD4 | OKT4 | Alexa 488 | BioLegend | 317420 |
| CD8 | SK1 | Alexa 700 | BioLegend | 344724 |
| OX-40 (CD134) | ACT35 | PE-Dazzle 594 | BioLegend | 350020 |
| CD69 | FN-50 | PE | BioLegend | 310906 |
| 4-1BB (CD137) | 4B4-1 | BV421 | BioLegend | 309820 |
| CD14 | HCD14 | PerCP-Cy5.5 | BioLegend | 325622 |
| CD16 | B73.1 | PerCP-Cy5.5 | BioLegend | 360712 |
| CD19 | HIB19 | PerCP-Cy5.5 | BioLegend | 302230 |
| Live/dead dye | | eFluor 506 | Invitrogen | 65-0866-14 |
| **Intracellular cytokine stimulation (ICS)** | | | | |
| **Antibody target** | **Clone** | **Fluorophore** | **Vendor** | **Catalog #** |
| CD4 | A161A1 | PE | BioLegend | 357404 |
| CD8 | SK1 | Alexa 700 | BioLegend | 344724 |
| CD14 | HCD14 | PerCP-Cy5.5 | BioLegend | 325622 |
| CD16 | B73.1 | PerCP-Cy5.5 | BioLegend | 360712 |
| CD19 | HIB19 | PerCP-Cy5.5 | BioLegend | 302230 |
| IFN$\gamma$ | 4S.B3 | Alexa 647 | BioLegend | 502516 |
| TNF$\alpha$ | MAb11 | Alexa 488 | BioLegend | 502915 |
| IL-2 | MQ1-17H12 | BV421 | BioLegend | 500328 |
| IL-4 | MP4-25D2 | APC-Cy7 | BioLegend | 500833 |
| IL-10 | JES3-9D7 | PE-Dazzle 594 | BioLegend | 501426 |
| Live/dead dye | | eFluor 506 | Invitrogen | 65-0866-14 |
| **Tetramer panel** | | | | |
| **Antibody target** | **Clone** | **Fluorophore** | **Vendor** | **Catalog #** |
| CD8 | SK1 | PE-Cy7 | eBioscience | 25-0087-42 |
| CD4 | RPA-T4 | PerCP-Cy5.5 | BioLegend | 300530 |
| CD14 | HCD14 | PerCP-Cy5.5 | BioLegend | 325622 |
| CD16 | B73.1 | PerCP-Cy5.5 | BioLegend | 360712 |
| CD19 | HIB19 | PerCP-Cy5.5 | BioLegend | 302230 |
| CCR7 | G043H7 | Alexa 488 | BioLegend | 353206 |
| CD45RA | HI100 | APC/Fire 750 | BioLegend | 304152 |
| Streptavidin | | PE | Invitrogen | S866 |
| Streptavidin | | APC | Invitrogen | S868 |
| Streptavidin | | BV421 | BioLegend | 405225 |
| Streptavidin | | PE-Dazzle 594 | BioLegend | 405247 |
| Live/dead dye | | eFluor 506 | Invitrogen | 65-0866-14 |

**Supplemental Table 1. Flow cytometry panel overview.** Antibodies labeled with the same fluorophore in a given panel were used to a create a ‘dump’ channel for exclusion of the designated populations.

| Tetramer panel 1 | | | |
|---|---|---|---|
| **Epitope** | **MHC I restriction** | **Fluorophore 1** | **Fluorophore 2** |
| YLQPRTFLL | A*02:01 | PE | APC |
| RLQSLQTYV | A*02:01 | PE | BV421 |
| VVFLHVTYV | A*02:01 | PE | PE-Dazzle 594 |
| SPRRARSVA | B*07:02 | PE-Dazzle 594 | BV421 |
| APHGVVFL | B*07:02 | PE-Dazzle 594 | APC |
| **Tetramer panel 2** | | | |
| **Epitope** | **MHC I restriction** | **Fluorophore 1** | **Fluorophore 2** |
| KCYGVSPTK | A*03:01 | PE | APC |
| GVYFASTEK | A*03:01 | PE | BV421 |
| GVYFASTEK | A*11:01 | PE | PE-Dazzle 594 |
| GTHWFVTQR | A*11:01 | PE-Dazzle 594 | BV421 |
| RLFRKSNLK | A*11:01 | PE-Dazzle 594 | APC |
| **Tetramer panel 3** | | | |
| **Epitope** | **MHC I restriction** | **Fluorophore 1** | **Fluorophore 2** |
| KCYGVSPTK | A*03:01 | PE | APC |
| GVYFASTEK | A*03:01 | PE | BV421 |
| LTDEMIAQY | A*01:01 | PE | PE-Dazzle 594 |

**Supplemental Table 2**. **Overview of pMHC I tetramer panels.** The epitope, MHC I restriction, and fluorophore combinations used for the indicated tetramer panels are shown.

| Sample ID | Treatment | HLA restriction for tetramer staining | | | Tetramer panel |
|---|---|---|---|---|---|
| HCCOV001 | HC | HLA-A*11:01 | | | 2 |
| HCCOV003 | HC | HLA-A*03:01 | HLA-A*11:01 | | 2 |
| HCCOV009 | HC | HLA-A*11:01 | | | 2 |
| HCCOV011 | HC | HLA-A*02:01 | HLA-B*07:02 | | 1 |
| HCCOV015 | HC | HLA-A*02:01 | | | 1 |
| MSCOV020 | None | HLA-A*03:01 | | | 2 |
| MSCOV027 | None | HLA-A*02:01 | HLA-B*07:02 | | 1 |
| MSCOV059 | None | HLA-A*02:01 | | | 1 |
| MSCOV070 | None | HLA-A*02:01 | | | 1 |
| MSCOV082 | None | HLA-A*03:01 | | | 2 |
| MSCOV009 | GA | HLA-B*07:02 | | | 1 |
| MSCOV047 | GA | HLA-B*07:02 | | | 1 |
| MSCOV077 | GA | HLA-B*07:02 | HLA-A*11:01 | | 1 & 2 |
| MSCOV010 | DMF | HLA-A*01:01 | | | 3 |
| MSCOV021 | DMF | HLA-A*03:01 | | | 2 |
| MSCOV035 | DMF | HLA-A*03:01 | HLA-A*11:01 | | 2 |
| MSCOV038 | DMF | HLA-A*03:01 | HLA-A*01:01 | | 3 |
| MSCOV089 | DMF | HLA-B*07:02 | HLA-A*03:01 | | 1 & 2 |
| MSCOV037 | NTZ | HLA-A*03:01 | HLA-A*01:01 | | 3 |
| MSCOV039 | NTZ | HLA-A*02:01 | | | |
| MSCOV052 | NTZ | HLA-A*02:01 | HLA-B*07:02 | HLA-A*03:01 | 1 & 2 |
| MSCOV057 | NTZ | HLA-A*03:01 | | | 2 |
| MSCOV069 | NTZ | HLA-B*07:02 | | | 1 |
| MSCOV016 | S1P | HLA-A*02:01 | | | 1 |
| MSCOV050 | S1P | HLA-B*07:02 | HLA-A*03:01 | | 1 & 2 |
| MSCOV060 | S1P | HLA-A*02:01 | HLA-B*07:02 | | 1 |
| MSCOV072 | S1P | HLA-A*01:01 | | | 3 |
| MSCOV012 | RTX | HLA-A*01:01 | | | 3 |
| MSCOV025 | RTX | HLA-B*07:02 | | | 2 |
| MSCOV049 | RTX | HLA-A*11:01 | | | 2 |
| MSCOV074 | RTX | HLA-A*02:01 | | | 1 |
| MSCOV076 | RTX | HLA-A*03:01 | HLA-A*11:01 | | 2 |
| MSCOV084 | RTX | HLA-A*02:01 | | | 1 |
| MSCOV014 | OCR | HLA-A*11:01 | | | 2 |
| MSCOV022 | OCR | HLA-B*07:02 | | | 1 |
| MSCOV045 | OCR | HLA-B*07:02 | | | 1 |
| MSCOV056 | OCR | HLA-A*02:01 | | | 1 |
| MSCOV058 | OCR | HLA-A*03:01 | | | 2 |
| MSCOV064 | OCR | HLA-A*03:01 | HLA-A*01:01 | | 3 |

**Supplemental Table 3.** Overview of patient samples used for pMHC I tetramer analysis. HLA restriction refers to the relevant MHC I alleles used for tetramer staining. The tetramer panel refers to the panels outlined in Supplemental Table 2.

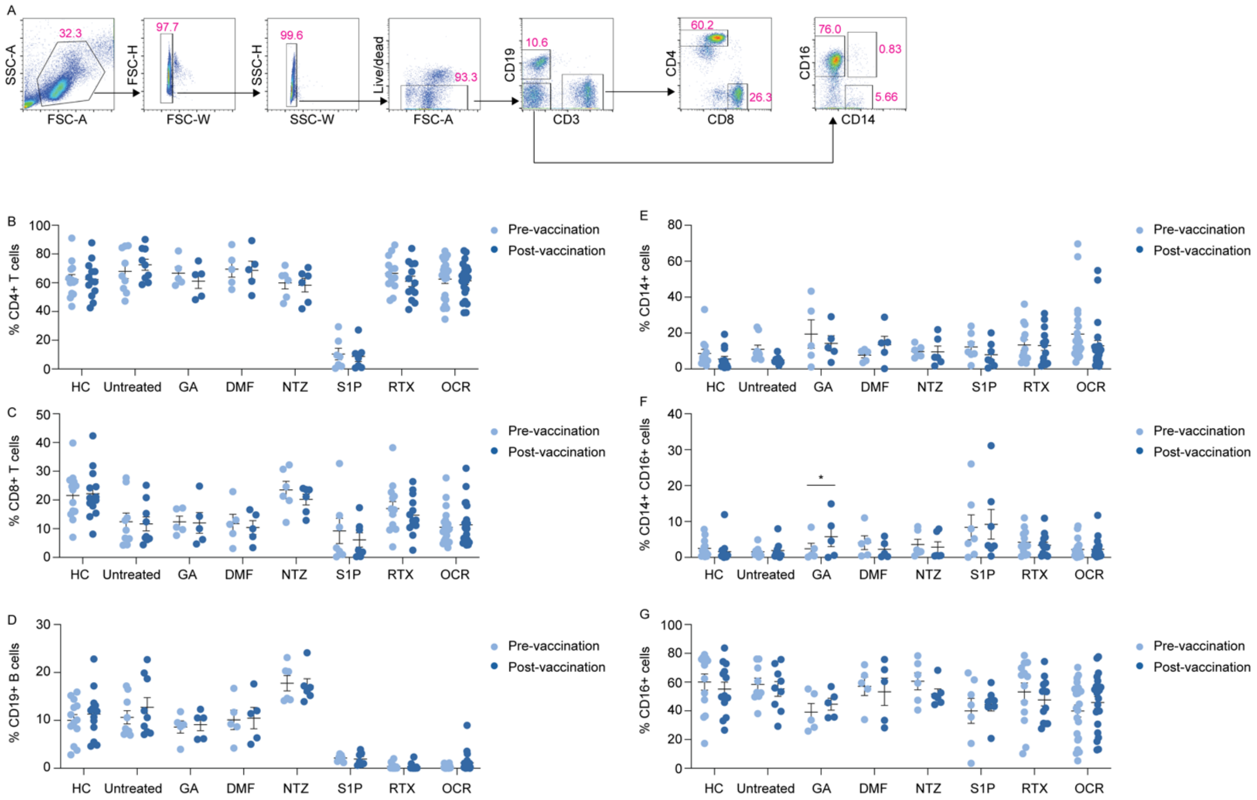


**Supplemental Figure 1.** Representative flow cytometry analysis for identification of immune cell subsets (**A**). Overview of immune cell subsets across all cohorts before and after vaccination: CD4+ T cells (**B**), CD8+ T cells (**C**), CD19+ B cells (**D**), CD14+ cells (**E**), CD14+ CD16+ cells (**F**), CD16+ cells (**G**). Comparisons of pre- and post-vaccination responses by multiple paired t-tests and comparisons of different patient cohorts by 2-way ANOVA with multiple comparisons.

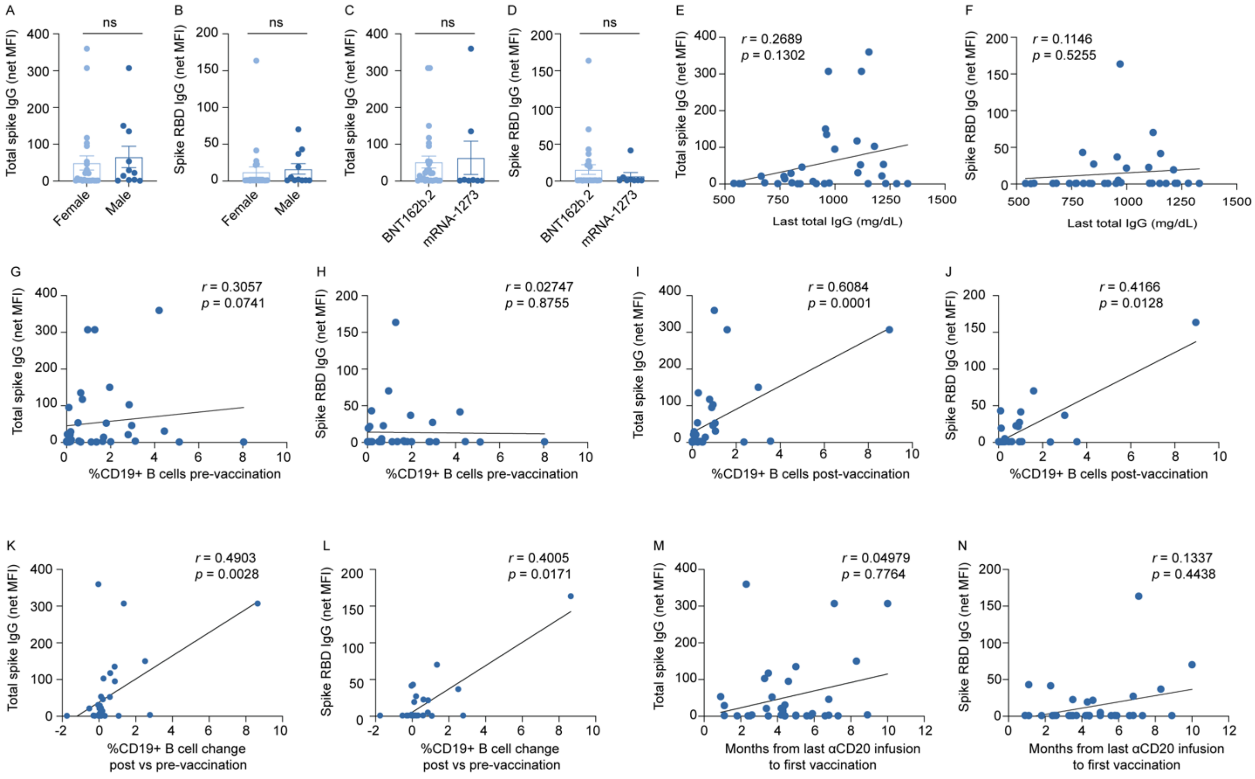


**Supplemental Figure 2. Analysis of vaccine-elicited spike-specific antibody responses in anti-CD20 mAb-treated MS patients.** Post-vaccination net MFI of total spike IgG (**A, C**) and spike RBD IgG (**B, D**) by gender (**A-B**) and mRNA vaccine type (**C-D**), comparisons by Mann-Whitney. Net MFI of total spike IgG and spike RBD IgG versus last total IgG (**E-F**), %CD19+ B cells pre-vaccination (**G-H**), %CD19+ B cells post-vaccination (**I-J**), %CD19+ B cell change pre- vs post-vaccination (**K-L**), and interval (months) from last anti-CD20 mAb infusion to first vaccination (**M-N**), measured by simple linear regression and Spearman correlation.

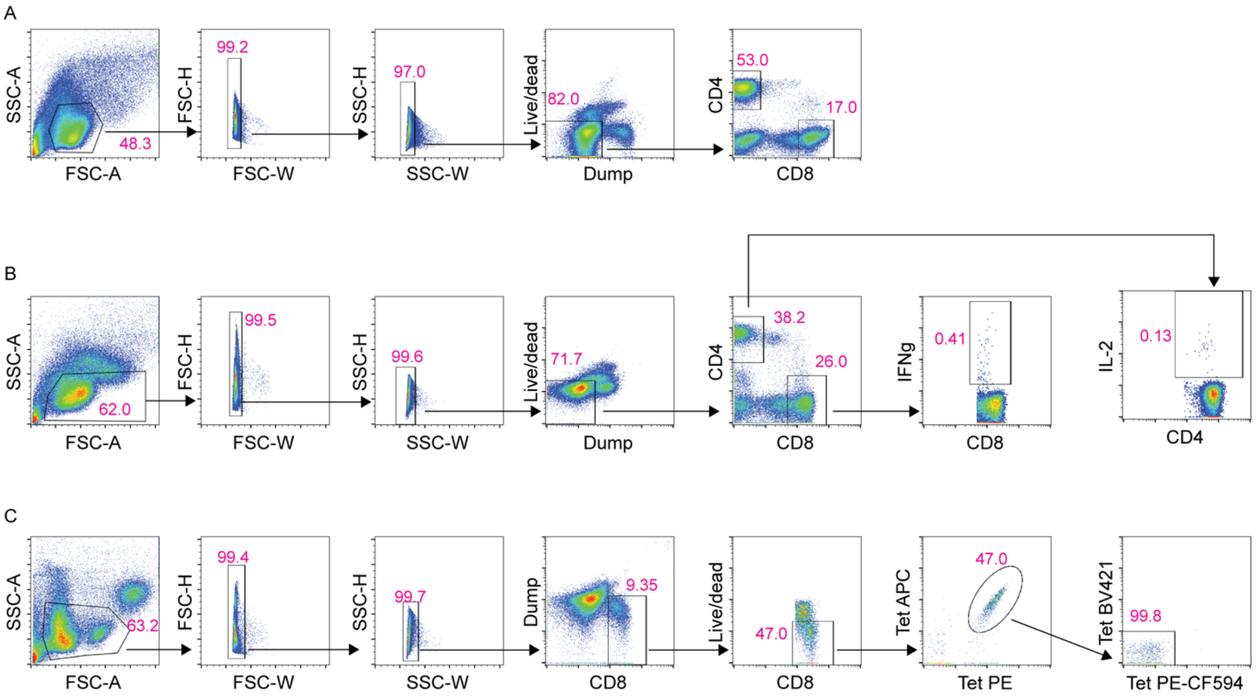


**Supplemental Figure 3. Gating overview for flow cytometry analysis.** Representative gating strategies for AIM (**A**), ICS (**B**), and pMHC I tetramer enrichment (**C**). T cells were identified by live single cell lymphocytes that were dump antibody negative using the antibody panels from Supplemental Table 1. For tetramer enrichment, CD8+ T cells that were tetramer-positive in two fluorophores were subsequently gated on cells negative for the remaining two fluorophores to ensure specificity of tetramer binding.
